# Association of sexual orientation with substance use in Colombian adults experiencing homelessness

**DOI:** 10.1101/2024.06.02.24308344

**Authors:** Adalberto Campo-Arias, Carlos Alejandro Pineda-Roa, Edwin Herazo

## Abstract

The study’s objective was to quantify the association of sexual orientation with substance use among Colombian adults experiencing homelessness. A secondary analysis of the Census of Street Inhabitants - CHC-2020 was completed. Three thousand five hundred forty-seven participants between 18 and 75 years participated, and 3.16% reported lesbian, gay, or bisexual (LGB) sexual orientation. LGB sexual orientation was associated with alcohol consumption (OR = 2.14, 95%CI 1.46 - 3.14]), cigarettes (OR = 1.70, 95%CI 1.12 - 2.56), cannabis (OR = 1.58, 95%CI 1.07 - 2.33), cocaine (OR = 2.13, 95%CI 1.30 - 3.50) and benzodiazepines (OR = 2.15, 95%CI 1.19-3.89). In conclusion, Colombian adults experiencing homelessness with gay or lesbian sexual orientation have twice the risk of consuming alcohol, cigarettes, cannabis, cocaine, and benzodiazepines than heterosexuals. More studies are needed on this population.

---

The homelessness situation represents a syndemic, given that various adversities and risks converge (Mabhala et al., 2021; Singer & Clair, 2003). People experiencing homelessness show a high frequency of health risk behaviors (Ecker et al., 2019; Oppenheimer et al., 2016). Substance use is highly prevalent in people experiencing homelessness and is associated with proximal (demographic characteristics) and distal (political, legal, or cultural) factors or determinants (Ecker et al., 2019; Flike & Aronowitz, 2021).

In the general adult population, lesbian, gay, or bisexual (LGB) sexual orientation has been significantly related to a higher frequency of substance use in several studies. For example, Kahle et al. (2020) found that LGB people were 50% more likely to have substance use disorders than heterosexual people. Likewise, Li et al. (2021), in a meta-analysis, found higher prevalences of cigarette smoking in LGB people than in heterosexuals: 37.7% in bisexual women; 31.7%, lesbians; 30.5%, homosexual men; 30.1%, bisexual men (30.1%); 21.0%, heterosexual men; and 16.6% heterosexual women.

However, the association of LGB sexual orientation and substance use in people experiencing homelessness is inconsistent in the few available studies that included young adults; there are no adult studies in a wide age range. In adolescents and young adults, Santa Maria et al. (2018) documented that the use of synthetic cannabis, stimulants, and opioids was significantly higher in LGB participants. However, Flentje et al. (2016) and Tyler and Ray (2019) highlighted that substance use was similar among adolescents and young adults experiencing homelessness regardless of sexual orientation.

LGB people are at high risk for experiencing homelessness (Corliss et al., 2011; Lim et al., 2023; McCarthy & Parr, 2022). In turn, homelessness increases the probability of substance abuse in LGB populations (Lim et al., 2023; Rosario et al., 2012). Furthermore, substance use deteriorates, even more, the quality of life of people experiencing homelessness, regardless of sexual orientation or age (Flike & Aronowitz, 2021), and increases the possibility of discrimination (Zerger et al., 2014).

It must be borne in mind that discrimination is a significant stressor for people living in social exclusion. People experiencing homelessness are frequently discriminated against (Vásquez et al., 2021). In addition, Meyer (2003), in minority stress theory, proposed that LGB people face negative situations related to sexual orientation and are at greater risk of stress and substance use.

The present study explores the relationship between sexual orientation and substance use in adults experiencing homelessness of a wide range of ages. To date, little has been studied in low- and middle-income countries. Colombia is a country with upper middle income, high inequality, and poverty (Chacón & Ramírez, 2020), Christian religious beliefs, and, therefore, high heteronormativity (Palacio, 2017). These characteristics are related to the attitude toward sexual orientations and, consequently, are confounding variables in the association between sexual orientation and substance use, even among people experiencing homelessness (Ecker et al., 2019; Flike & Aronowitz, 2021).

This information may be relevant to the Colombian healthcare system, which favors differential care in all contexts to guarantee the full enjoyment of the right to health for all citizens (García-Balaguera, 2018).

The study’s objective was to quantify the association of sexual orientation with substance use in Colombian adults experiencing homelessness.

## METHOD

### Design

This study analyzes secondary data from the Homeless Inhabitants Census –HIC– 2020 carried out by the National Administrative Department of Statistics of Colombia—DANE, identified with the code DANE-DCD-CHC-2020, available anonymized on the institution’s page (DANE, 2021).

The HIC-2020 census study considers 31 of the 32 Colombian departments (states) and 661 of the 1,103 Colombian municipalities in 2020. Participants over 18 years of age were included.

### Measurements

The interview included demographic information: Age, gender, level of education attained, ethnic-racial self-recognition, and homelessness time. The current alcohol, cigarettes, cannabis, inhalants, cocaine, cocaine base, heroin, benzodiazepines, and other substances were asked.

All variables were divided into two categories. Age classified as emergent adults (between 18 and 29 years) and post-emergent adults (30 years and over). The level of education was divided into middle-school grades or less and high-school grades or more. The racial-ethnic self-recognition was classified into minorities (Indigenous, gypsy, or Afro-Colombian) and another group (not identified with the previous ones). The time experiencing homelessness was worked with two categories with a cut-off point of five years: less than five years and more than five years. Current substance use followed the yes or no response pattern.

Sexual orientation was investigated with the question. Do you recognize yourself in your sexual orientation? Five options were recorded: Heterosexual? Homosexual? Bisexual? Do not know? And No answer? This category was re-categorized into three groups: Heterosexual, GLB, and does not know or respond.

### Statistical Analysis

Descriptive analysis included the data distribution, mean and standard deviation for age and frequencies, and percentages for categorical variables. Sexual orientation was taken as an independent variable, and substance use was taken as a dependent variable. The remaining measurements were considered covariates or confounding variables. Finally, they established crude and adjusted odds ratios (OR) and 95% confidence intervals (95% CI). The analysis was carried out with the IBM-SPSS program, version 27.

## RESULTS

Four thousand nine hundred seventy-seven people experiencing homelessness were surveyed. People experiencing homelessness were counted in 283 municipalities. Three hundred seventy-one municipalities did not have certified homelessness situations, and in 7 municipalities, adverse situations prevented the census.

Of the 4,109 562 people (13.68%) who did not report sexual orientation were excluded from the analysis. The participants who omitted sexual orientation were more frequently women, post-emergent age, high-school education or higher, ethnic-racial minority, and five years or more of experiencing homelessness.

The ages of the participants (N = 3,547) were observed between 18 and 78 years (M = 40.61, SD = 15.18); 86.41% (n = 3,065) men; 63.35% (n = 2,247) with lower secondary education less than schooling, 7.56% (n = 268) self-recognized as ethnic minorities; 37.61% (n = 1,334) with five years or less of experiencing homelessness and 3.16% (n = 112) reported LGB sexual orientation. The current frequency of substance use is reported in Table 1.

**Table 1.** Frequency of current substance use (N = 3,547).

| Substance | n (%) |
| --- | --- |
| Alcohol | 1,078 (30.4) |
| Benzodiazepines | 236 (6.7) |
| Cigarette | 2,085 (58.8) |
| Cannabis | 1,763 (49.7) |
| Cocaine | 398 (11.2) |
| Cocaine base | 1,993 (56.2) |
| Heroin | 249 (7.0) |
| Inhalants | 474 (13.5) |

The crude association of diverse sexual orientation was significant for the substances studied, except the consumption of cocaine base and heroin. After adjusting for age, gender, and education (confusion variables), the association with inhalant use lost statistical significance. See crude and adjusted associations in Table 2.

**Table 2.** Associations of diverse sexual orientation with current substance use (crude and adjusted for age, gender, and education).

| Substance | OR (95% CI) | aOR (95% CI) |
| --- | --- | --- |
| Alcohol | 2.04 (1.39 – 2.97) | 2.14 (1.46 – 3.14) |
| Cigarette | 1.56 (1.04 – 2.35) | 1.70 (1.12 – 2.56) |
| Cannabis | 1.47 (1.01 – 2.15) | 1.58 (1.07 – 2.33) |
| Inhalants | 1.61 (1.01 – 2.60) | 1.57 (0.96 – 2.55) |
| Cocaine | 1.87 (1.15 – 3.04) | 2.13 (1.30 – 3.50) |
| Cocaine base | 1.36 (0.92 – 2.01) | 1.35 (0.90 – 2.01) |
| Heroin | 1.16 (0.58 – 2.33) | 1.17 (0.58 – 2.39) |
| Benzodiazepines | 2.07 (1.16 – 3.68) | 2.15 (1.19 – 3.89) |

a, adjusted.

## DISCUSSION

In a country of Christian religious affiliation and high heteronormativity, LGB sexual orientations are associated with the current consumption of alcohol, cigarettes, cannabis, cocaine, and benzodiazepines in a census sample of Colombian adults experiencing homelessness.

Few studies have explored the association of sexual orientation with substance use in adults experiencing homelessness. The available studies have been conducted with adolescents and young adults, which limits comparisons with older age groups. The current findings are consistent with what was observed in the United States of America. Santa Maria et al. (2018), in 416 participants aged 16-24 years, 24% of LGB people, observed that LGB participants reported greater use of synthetic cannabis, stimulants, and opioids during the most recent month than their heterosexual counterparts. Consumption of alcohol and traditional cannabis was similar in both groups.

On the other hand, Flentje et al. (2016), in 1,019 young adults, 639 men (2.50% gay or bisexual) and 372 women (8.87% lesbian or bisexual) documented that the consumption of alcohol and other substances was unrelated to sexual orientation. Tyler and Ray (2019), in a sample of 322 young people between 16 and 26 years old, 21% of LGB participants, found that binge drinking, cannabis use, and other illegal substances were similar in LGB and heterosexual participants. The differences in the social and cultural characteristics of the participants could account for the divergence in the findings to date (Grimes & Schulz, 2002).

As suggested by the minority stress theory, sexually diverse people face a more significant number of particular psychosocial stressors due to sexual orientation and present greater vulnerability to stress and substance use (Meyer, 2003). The relationship of diverse sexual orientation with substance use may indicate that the negative social connotations of sexual orientation represent another stressor for the population experiencing homelessness (Meyer, 2003). This convergence is a clear example of intersectoral discrimination in which negative connotations for LGB sexual orientation and homelessness are added (Vásquez et al., 2021).

### Practical Implications

The association of sexual orientation with substance use in people experiencing homelessness is relevant for public health. These variables interact in complex ways. Homelessness situation increases the probability of substance use (Rosario et al., 2012). LGB sexual orientation increases the risk of experiencing homelessness and substance use (Corliss et al., 2011; Marshall et al., 2008). Furthermore, finally, substance use is a predictor of homelessness situation (McVicar et al., 2015).

Moreover, homelessness situation, LGB sexual orientation, and substance use are three different psychosocial stressors (Meyer, 2003). This grouping configures a circumstance of intersectoral stigma-discrimination (Vásquez et al., 2021) and is an independent variable associated with suicidal behaviors (Hawton & van Heeringen, 2009).

People experiencing homelessness represent approximately 0.01% of the population of Colombia, a country of around 50 million inhabitants (DANE, 2021). They are an orphan social group marked by intersectoral stigma-discrimination (Vásquez et al., 2021). Official institutions must break down access barriers to meet this group’s health needs in a differentiated way and consider the challenges posed by the post-conflict context, the reactivation of armed violence, and the post-pandemic coronavirus disease (García-Balaguera, 2018).

### Study strengths and limitations

One of the strengths of this study was having a national census sample. In addition, it is one of the first investigations to explore the association of sexual orientation and substance use in people experiencing homelessness in a middle-income country with high social inequalities indicators. However, the study has the limitation that the participants who omitted sexual orientation had statistically different demographic characteristics, which prevents generalizations; furthermore, how homelessness situation and LGB sexual orientation were measured.

### Conclusions

LGB sexual orientation presents twice the risk of alcohol, cigarette, cannabis, cocaine, and benzodiazepine consumption in Colombian adults experiencing homelessness than their heterosexual counterparts. More studies are needed on this population to know and differentially meet the needs of this group in the context of the post-Colombian armed conflict and the post-pandemic caused by coronavirus disease.

## Data Availability

The data supporting this study’s findings are available at http://microdatos.dane.gov.co/index.php/catalog/703.

